# Unveiling Endoglin non canonical regulation: spotlight on the new role of the uPAR pathway

**DOI:** 10.1101/2024.01.28.24301864

**Authors:** Gaëlle Munsch, Carole Proust, Clémence Deiber, Caroline Meguerditchian, Ilana Caro, Maud Tusseau, Alexandre Guilhem, Shirine Mohamed, INVENT consortium, Aurélie Goyenvalle, Stéphanie Debette, Béatrice Jaspard-Vinassa, Sophie Dupuis-Girod, David-Alexandre Trégouët, Omar Soukarieh

## Abstract

Endoglin, encoded by *ENG*, is a transmembrane glycoprotein crucial for endothelial cell biology. Loss-of-function *ENG* variants cause Hereditary Hemorrhagic Telangiectasia (HHT). Despite advances in HHT diagnosis and management, the molecular origin of some cases and the source of clinical heterogeneity remain unclear.

We propose a comprehensive *in silico* analysis of all 5’UTR *ENG* single nucleotide variants that could lead to Endoglin deficiency by altering upstream Open Reading Frames (upORFs). Experimentally, we confirm that variants creating uAUG-initiated overlapping upORFs associate with reduced Endoglin levels *in vitro* and characterize the effect of a uCUG-creating variant identified in two suspected HHT patients.

Using plasma proteogenomics resources, we identify eight loci associated with soluble Endoglin levels, including *ABO* and uPAR-pathway loci and experimentally demonstrate the association between uPAR and Endoglin in endothelial cells.

This study provides new insights into Endoglin’s molecular determinants, opening avenues for improved HHT management and other diseases involving Endoglin.

**Key points:**

- New insights on the characterization of *ENG* non-coding variants, in particular those altering upstream Open Reading Frames in the 5’UTR.
- Leverage of large-scale plasma proteogenomics results combined with functional assays revealed new actors in Endoglin regulation.

## Introduction

Endoglin is a transmembrane glycoprotein that plays a major role in endothelial cell biology and is highly involved in angiogenesis^1,2^. Endoglin has mainly been known as co-receptor in response to bone morphogenetic proteins belonging to the transforming growth factor β (TGF-β) superfamily^3–5^. Loss-of-function (LoF) mutations in the *ENG* gene, which codes for Endoglin, are responsible for Hereditary Hemorrhagic Telangiectasia (HHT), a multiorgan rare vascular disease^6^.

The clinical diagnosis of HHT is based on Curaçao criteria which include family history, epistaxis, multiple telangiectasias, and visceral vascular lesions such as gastrointestinal telangiectasia and/or arteriovenous malformations (AVMs)^7^. Depending on the identified criteria in patients, the clinical diagnosis of HHT can be classified as definite, suspected, or unlikely. Tremendous efforts have been made in order to identify new genetic drivers^8^ and molecular explanations at the origin of HHT^9,10^. However, around 10% of cases are still with unclear molecular origin^6,8^. In addition, there is a large inter-individual variability in the clinical manifestations and complications of HHT, even in patients carrying the same pathogenic *ENG* mutations^11,12^. This variability requires a multidisciplinary approach for both clinical diagnosis and treatment ^11,12^. In consequence, resolving unexplained cases and bringing new elements to better understand phenotype heterogeneity is necessary to ameliorate the molecular diagnosis and management of HHT.

For a long time, the search for pathogenic *ENG* mutations in HHT patients was mainly restricted to exonic/flanking intronic regions^6,10,13^. However, we and others have recently reported 5 pathogenic variations in the 5’UTR of *ENG* identified in HHT patients with definite diagnosis^14–19^. Acting as LoF mutations and associating with Endoglin deficiency^14^, these variations create upstream AUGs (uAUGs) in frame with the same stop codon in position c.125 (uStop-c.125) and resulting in overlapping upstream Open Reading Frames (uoORFs). These data suggested that *ENG* could be enriched in HHT-causing uAUG-creating variants. Given the growing evidence that non-canonical translation initiation sites (TIS) differing by one nucleotide from the AUG codon can also initiate translation^20^, and despite such evidence in the context of HHT, we hypothesize that upstream ORFs (upORFs) starting with non-canonical TIS could also contribute to regulate the translation of the Endoglin coding sequence (CDS). To provide a comprehensive overview of genetic variations altering upORFs in the 5’UTR of *ENG* and to improve molecular diagnosis of HHT, we here use an updated version of the bioinformatics tool MORFEE^21^ that now annotates any single nucleotide variations (SNVs) that could create canonical and non-canonical uTIS, but also those creating new stop codons, and/or deleting existing upstream stop codons (uStop). Thanks to MORFEE, we then perform an exhaustive bioinformatics analysis of all possibleSNVs in the 5’UTR of *ENG* that could alter upORFs. The functional impact on Endoglin levels of a selection of variants was assessed.

The pathophysiology of HHT and the molecular pathways regulating Endoglin numerous functions, in particular, are not yet completely understood. Different studies have highlighted the role of transmembrane Endoglin in leukocyte adhesion and transmigration under inflammatory conditions and cancer-associated fibroblasts^22,23^, in addition of its role in the TGF-β signaling pathway. Moreover, the soluble form of Endoglin (sol-ENG), cleaved from the membrane by matrix metalloproteases MMP12 and MMP14, has also been studied recently and has been described to be implicated in platelets biology^24^. Interestingly, HHT patients present reduced sol-ENG levels in plasma, which has been proposed as a biomarker in HHT^25,26^. In parallel, sol-ENG reduced the number of AVMs, most severe complication in HHT, in a HHT mouse model. This suggests an anti-angiogenic role of sol-ENG antagonistic of transmembrane Endoglin function^27^.

To identify new regulators of *ENG* that could subsequently contribute to a better understanding of the clinical heterogeneity of HHT, we additionally leveraged results from large-scale plasma proteogenomics resources that measured plasma Endoglin levels. This approach is based on the hypothesis that any biological factors involved in the regulation of Sol-ENG may reveal novel elements contributing to Endoglin function in general, and HHT in particular. Furthermore, Endoglin and its soluble form have been shown to be involved in many human diseases beyond HHT^28^ such as thrombosis^29^, coronary atherosclerosis^30^, preeclampsia^31^, hypertension^32^, auto-immune disease^33^, and some cancers^34^. As a consequence, dissecting the genetic regulation of Endoglin holds the potential not only to enhance molecular diagnosis and clinical management for HHT patients but also to deepen understanding of the pathophysiology underlying more common diseases.

## Results

### Identification of all possible upORF-altering variations in the 5’UTR of *ENG*

In total, 909 possible SNVs in the 5’UTR of *ENG* resulted from the *in silico* mutational saturation (Supplemental Table 1). MORFEE annotated 328 of them as creating uTIS, uStop, and/or to delete existing uStop. These annotated variations can be at the origin of 360 upORFs (Figure 1a, Supplemental Table 2). More precisely, 255 variants are predicted to create new uTIS, 30 to create new uStop and 12 to delete existing uStop (Figure 1a). The remaining 31 SNVs show multiple consequences (Figure 1a; Supplemental Table 3). While the majority of annotated variants do not exist in databases and correspond to artificial ones, 18 of them, all creating uTIS, are reported in ClinVar as candidates for HHT. Of these 18 variants, 13 are classified as variants of unknown significance (VUS) or with conflicting interpretations (Supplemental Table 4). In addition, 14 additional uTIS-creating variants have been reported in GnomAD V4 0.0 database with minor allele frequency lower than 0.01% and without any evidence of association with HHT (Supplemental Table 5).

**Figure 1.**
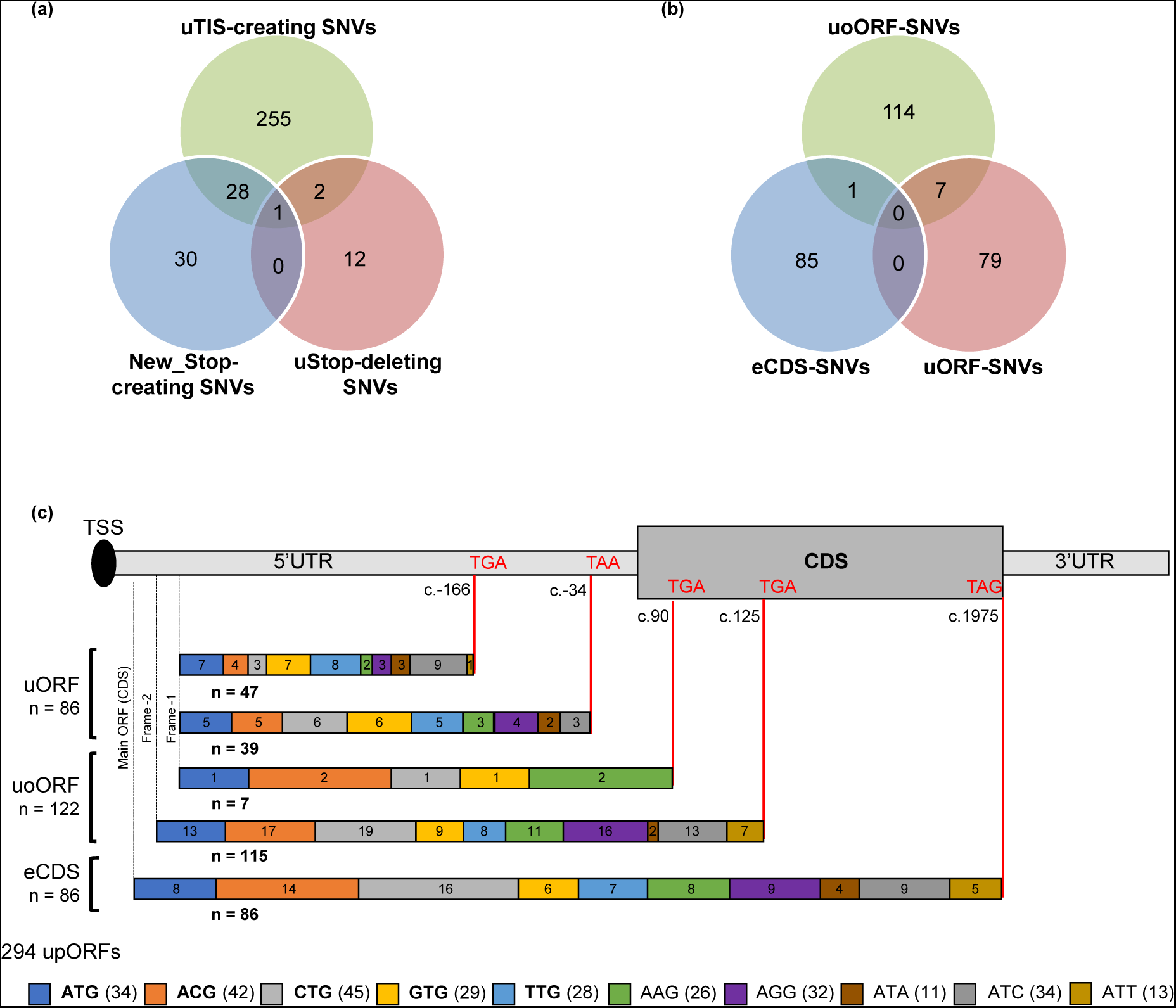
Identification of all possible single nucleotide variants (SNVs) modifying or creating upstream open reading frames (upORFs) in the 5’UTR of ENG. (a) MORFEE annotated SNVs that modify existing upORFs or create new ones (New_Stop-creating, uStop-deleting SNVs and uTIS-creating SNVs). (b) Three types of upORFs can result from the creation of upstream translation initiation sites (uTIS) in the 5’UTR of ENG. uoORF-SNVs, variants at the origin of upORFs overlapping the coding sequence (CDS); eCDS-SNVs, variants creating elongated CDS; uORF-SNVs, variants creating fully upstream upORFs. (c) Detailed illustration of upORFs generated by uTIS-creating SNVs. The type of uTIS and position of stop codons associated with the generated upORFs, as well as the number of each type of upORF and of uTIS are indicated. TSS, translation start site.

In more detail, 286 SNVs creating canonical or non-canonical uTIS could be at the origin of 294 upORFs, with the understanding that a given SNV may create different uTIS (Figure 1b; Supplemental Table 6). Among these upORFs, 86 are fully located in the 5’UTR (uORFs) ending at 2 different stop codons (c.- 166 and c.-34), 122 are overlapping with the CDS (uoORFs) ending with stop codons at position c.90 or c.125, and 86 correspond to elongated CDS (eCDS) ending at the main stop codon (i.e. natural stop codon of the CDS) (Figure 1c).

Interestingly, 8 upORFs have been reported in public databases (www.sORFs.org^35^ ; https://metamorf.hb.univ-amu.fr/^36^ ; https://vutr.rarediseasegenomics.org/ ; and https://smorfs.ddnetbio.com^37^) as naturally existing in the 5’UTR of *ENG*. Five of these upORFs end with the stop codon located at c.-166, 2 with the stop codon at position c.-33 and 1 is overlapping the CDS and ends at the uStop-c.125 (Supplemental Figure 1a). Interestingly, MORFEE annotated 8 variations as deleting the uStop codon located at position c.-166, and 7 that delete the uStop codon at position c.-34, thus elongating these existing upORFs into either longer fully upstream ORFs (uORFs) ending at positions c.-34 or into uoORFs ending at new stop codon c.90, respectively (Supplemental Figure 1b). By contrast, MORFEE identified 17 variations that could create new stop codons then shortening existing upORFs reported in databases and 42 that could create new stop codons at the origin of new upORFs (Supplemental Figure 1c). None of the annotated uStop-deleting or new stop-creating variants in the 5’UTR of *ENG* are reported in ClinVar. However, 2 of those creating new stop codons (c.-253C>A and c.-87C>T) have been reported as rare in GnomAD V4 0.0 database with the c.- 87C>T showing a double consequence (Supplemental Table 5).

### Most *ENG* SNVs creating uAUGs in frame with the uStop-c.125 drastically alter the protein levels

Recently, we have demonstrated that 5 *ENG* 5’UTR variants identified in HHT patients and creating uAUG-initiated uoORFs all ending at the uStop-c.125, were responsible of decreased protein levels^14^, revealing their pathogenicity (Table 1). To extrapolate whether these deleterious effects also hold for any other uAUG-creating SNVs at the origin of uoORFs ending at the uStop-c.125, we conducted the same experimental work^14^ on the remaining 8 variations predicted by the MORFEE *in silico* analysis (Figure 2a; Table 1). No decrease of protein levels was observed for the c.-287C>A and c.-37G>T variations (Figure 2b-c). While the c.-287C>A variant showed similar protein levels comparing to the wild-type (WT), the c.-37G>T tended to associate with an increase of *ENG* levels in our assay (Figure 2b-c; Supplemental Figure 2a). Interestingly, the c.-37G>T variant is predicted to simultaneously create a uAUG-initiated uoORF and to shorten an existing upORF (Supplemental Table 3). All other 6 variations were associated with a protein level lower than 40% compared to the wild-type construct. No significant difference was observed in our RT-qPCR experiments between mutants and WT (Supplemental Figure 2b).

**Figure 2.**
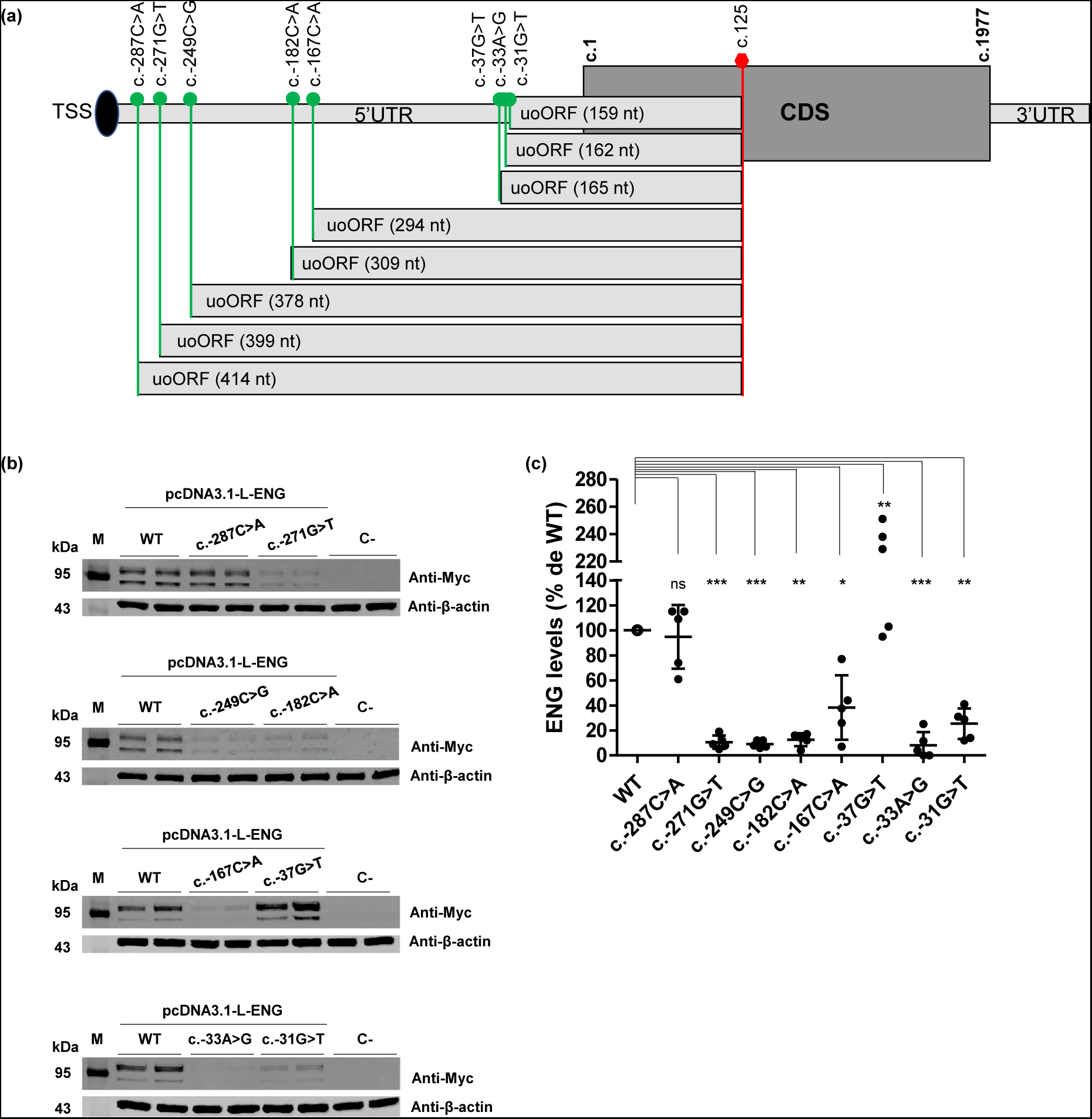
*ENG* variants at the origin of uAUGs in frame with the same stop codon at position c.125. (a) Eight variants in the 5’UTR create uAUG-initiated upstream overlapping open reading frames (uoORFs) with different sizes but ending at the same position. Position and identity of these variants and of created uAUG, size of the uoORFs, associated stop codon located at position c.125 and first and last positions of the CoDing Sequence (CDS) (c.1 and c.1977, respectively) are indicated. TSS, translation start site; CDS, CoDing Sequence. (b) Western blot results on total proteins extracted from transfected HeLa cells with 1 μg of pcDNA3.1-L-ENG constructs. Two bands of different molecular weights are observed for endoglin likely corresponding to more glycosylated (upper band) and less/non glycosylated (lower band) ENG monomers. Anti-Myc and anti-ENG correspond to the used antibodies for the target protein from HeLa and HUVECs, respectively, and anti-β-actin corresponds to the antibody used against the reference protein. kDa kilodalton, M protein ladder, WT wild type, C-negative control corresponding to pcDNA3.1-empty vector. Shown results are representative of 5 independent experiments. All blots were processed in parallel and derive from the same experiments. (c) Quantification of ENG steady-state levels in HeLa cells from (b). For quantification, the average of each duplicate has been calculated from the quantified values and ENG levels for each sample have been normalized to the corresponding β-actin levels then to the WT (%). The two bands obtained for the Endoglin, corresponding to the more glycosylated (upper band) and less/non glycosylated (lower band) ENG monomers 1, were taken together for the quantification. Graphs with standard error of the mean are representative of 5 independent experiments. ***, p-value < 10^-3^; **, p < 10^-2^; *, p < 5.10^-2^, ns, non-significant (two-factor ANOVA followed by Tukey’s multiple comparison test of variants versus WT).

**Table 1.** Characteristics of the 13 uAUGs and the studied uCUG created by variants in the 5’UTR of ENG. uTIS position in the L-ENG; NM_001114753.3 transcript (c.1 corresponds to the A of the main AUG) is indicated. The 5 variants from our previous study are bolded. Nucleotides at positions -3 and +4 relative to the predicted uTIS are in bold when corresponding to the most conserved nucleotide. The uTIS are underlined. Bolded scores are those higher than the predefined thresholds.

| uTIS-SNV | rsID | ClinVar | Protein levels *in vitro* | uoORF size (nt) | Kozak sequence | PreTIS score | TIS-Predictor score |
| --- | --- | --- | --- | --- | --- | --- | --- |
| c.-287C>A* | NA | No | ~WT | 414 | CAT<u>ATGC</u> | na | 0.42 |
| c.-271G>T* | NA | No | < 20% | 399 | **GCC**<u>ATGA</u> | na | **0.78** |
| c.-249C>G | NA | No | < 20% | 378 | **GCC**<u>ATGC</u> | na | **0.67** |
| c.-182C>A | NA | No | < 20% | 309 | TCC<u>ATGT</u> | **0.91** | 0.57 |
| c.-167C>A | NA | No | < 40% | 294 | CTC<u>ATGA</u> | **0.87** | 0.55 |
| **c.-142A>T** | NA | No | < 20% | 270 | CAG<u>ATGG</u> | **0.84** | **0.66** |
| **c.-127C>T** | rs1060501408 | Yes (P/LP) | < 20% | 255 | **GGG**<u>ATGC</u> | **0.84** | **0.69** |
| **c.-79C>T** | rs1564466502 | Yes (VUS) | < 20% | 207 | CCC<u>ATGC</u> | **0.67** | **0.65** |
| **c.-68G>A** | NA | No | < 50% | 195 | CCG<u>ATGC</u> | **0.89** | 0.61 |
| c.-37G>T* | NA | No | ≥ WT | 165 | CAC<u>ATGA</u> | **1** | 0.57 |
| c.-33A>G | NA | No | < 20% | 162 | **AGG**<u>ATGA</u> | **1** | **0.73** |
| c.-31G>T | NA | No | < 30% | 159 | **ATA**<u>ATGC</u> | **0.96** | 0,63 |
| **c.-10C>T** | rs756994701 | Yes (Conflicting) | < 40% | 138 | CCC<u>ATGT</u> | **0.85** | 0.61 |
| c.-76C>T | rs943786398 | Yes (VUS) | < 75% | 204 | **ACG**<u>CTGG</u> | **0.93** | **0.87** |

\*, in addition of the uAUG-creation, c.-287C>A and c.-37G>T are predicted to create new stop codons shortening existing upORFs, and c.-271G>T is predicted to simultaneously create a uAUA at the origin of a fully upstream upORF. nt, nucleotide; P, pathogenic; LP, Likely-Pathogenic; VUS, variant of unknown significance; na, non-applicable.

**Table 1. Characteristics of the 13 uAUGs and the studied uCUG created by variants in the 5'UTR of ENG.**
| uTIS-SNV | rsID | ClinVar | Protein levels *in vitro* | uoORF size (nt) | Kozak sequence | PreTIS score | TIS-Predictor score |
| --- | --- | --- | --- | --- | --- | --- | --- |
| c.-287C>A* | NA | No | ~WT | 414 | CAT<u>AT</u>GC | na | 0.42 |
| c.-271G>T* | NA | No | < 20% | 399 | **GCC**<u>AT</u>GA | na | **0.78** |
| c.-249C>G | NA | No | < 20% | 378 | **GCC**<u>AT</u>GC | na | **0.67** |
| c.-182C>A | NA | No | < 20% | 309 | TCC<u>AT</u>GT | **0.91** | 0.57 |
| c.-167C>A | NA | No | < 40% | 294 | CTC<u>AT</u>GA | **0.87** | 0.55 |
| **c.-142A>T** | NA | No | < 20% | 270 | CAG<u>AT</u>**GG** | **0.84** | **0.66** |
| **c.-127C>T** | rs1060501408 | Yes (P/LP) | < 20% | 255 | **GGG**<u>AT</u>GC | **0.84** | **0.69** |
| **c.-79C>T** | rs1564466502 | Yes (VUS) | < 20% | 207 | CCC<u>AT</u>GC | **0.67** | **0.65** |
| **c.-68G>A** | NA | No | < 50% | 195 | CCG<u>AT</u>GC | **0.89** | 0.61 |
| c.-37G>T* | NA | No | ≥ WT | 165 | CAC<u>AT</u>GA | **1** | 0.57 |
| c.-33A>G | NA | No | < 20% | 162 | **AGG**<u>AT</u>GA | **1** | **0.73** |
| c.-31G>T | NA | No | < 30% | 159 | **ATA**<u>AT</u>GC | **0.96** | 0,63 |
| **c.-10C>T** | rs756994701 | Yes (Conflicting) | < 40% | 138 | CCC<u>AT</u>GT | **0.85** | 0.61 |
| c.-76C>T | rs943786398 | Yes (VUS) | < 75% | 204 | **ACG**<u>CT</u>**GG** | **0.93** | **0.87** |

\*, in addition of the uAUG-creation, c.-287C>A and c.-37G>T are predicted to create new stop codons shortening existing upORFs, and c.-271G>T is predicted to simultaneously create a uAUA at the origin of a fully upstream upORF. nt, nucleotide; P, pathogenic; LP, Likely-Pathogenic; VUS, variant of unknown significance; na, non-applicable.

In total, 85% (11/13) of these variants (current and previous works^14^) decrease the ENG protein levels *in vitro*. The obtained results are partially consistent with bioinformatics predictions based ok Kozak sequence strength, TIS-predictor and PreTIS scores (Table 1, Supplemental results).

### New uoORF-creating variants identified in HHT patients

We used the generated catalog of variants annotated with MORFEE to retrospectively analyze *ENG* variants discovered in patients from the French National reference center for HHT with unresolved molecular diagnosis. We thus identified 2 uTIS-creating variants, the aforementioned c.-33A>G and the c.-76C>T variants. The first one, creating a uAUG predicted to generate a uoORF ending at the c.125 codon and never reported in public databases, was identified in a patient with definite HHT according to Curaçao criteria. Our experimental study (Figure 2b-c) provided strong argument for its pathogenicity. The second variant, creating a non-canonical TIS (uCUG), is also predicted to generate a uoORF ending at the c.125 codon (Figure 3a). This variant (rs943786398) was detected in 2 unrelated patients with suspected HHT (Supplemental Table 7) and has been classified as VUS in ClinVar. The proband in the first family had an atypical presentation for HHT with stroke and deep vein thrombosis associated with few telangiectasias. In the second family, the proband presented with pulmonary AVM and the father was an asymptomatic carrier of the variant. Following the same experimental workflow as above, we observed that the c.-76C>T variant was associated with decreased Endoglin levels of more than 25% in comparison with the WT (Figure 3b-c). No significant difference of ENG transcript amounts was observed between c.-76C>T and WT by RT-qPCR (Supplemental Figure 2c). Very interestingly, the uCUG created by *ENG* c.-76C>T is encompassed by a strong Kozak sequence and carries very high scores with TIS-predictor and PreTIS (Table 1). Five additional variants creating non-canonical uTIS in frame with the stop codon at position c.125 have been reported in ClinVar (Supplemental Table 4).

**Figure 3.**
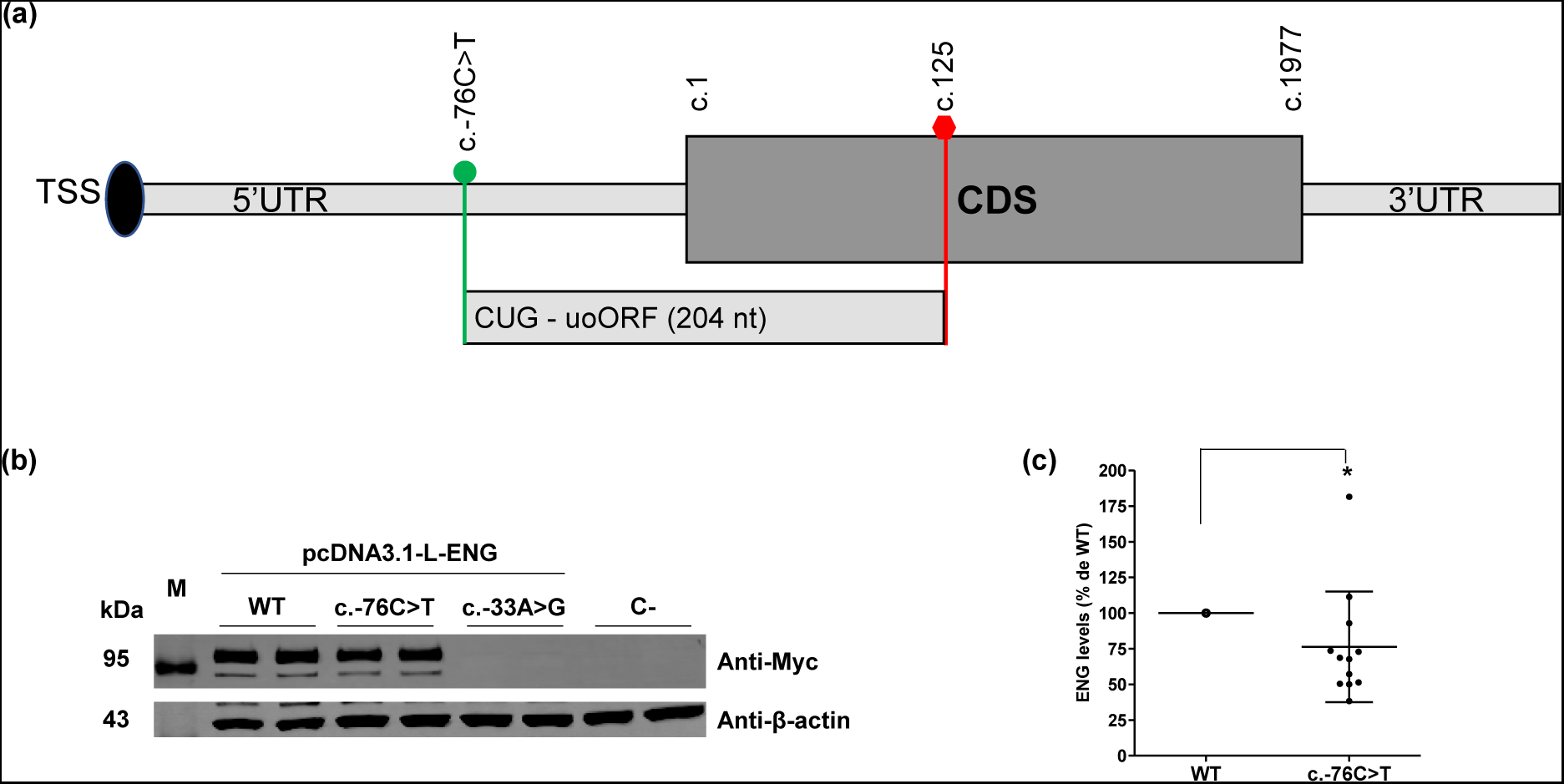
uoORF-creating variants identified in French HHT patient. (a) ENG c.-76C>T creating an upstream CUG in frame with the stop codon at position c.125 at the origin of an upstream Overlapping Open Reading Frame (uoORF) of 204 nucleotides (nt). Position and identity of the variant and of created uCUG, and first and last positions of the CoDing Sequence (CDS) (c.1 and c.1977, respectively) are indicated. (b) Western blot results on total proteins extracted from transfected HeLa cells with 1 μg of pcDNA3.1-L-ENG constructs. Two bands of different molecular weights are observed for endoglin likely corresponding to more glycosylated (upper band) and less/non glycosylated (lower band) ENG monomers. Anti-Myc and anti-ENG correspond to the used antibodies for the target protein from HeLa and HUVECs, respectively, and anti-β-actin corresponds to the antibody used against the reference protein. kDa kilodalton, M protein ladder, WT wild type, C-negative control corresponding to pcDNA3.1- empty vector. Shown results are representative of 5 independent experiments. All blots were processed in parallel and derive from the same experiments. (c) Quantification of ENG steady-state levels in HeLa cells from (b). For quantification, the average of each duplicate has been calculated from the quantified values and ENG levels for c.-76C>T sample have been normalized to the corresponding β-actin levels then to the WT (%). The two bands obtained for the Endoglin, corresponding to the more glycosylated (upper band) and less/non glycosylated (lower band) ENG monomers 1, were taken together for the quantification. Graphs with standard error of the mean are representative of 12 independent experiments. *, p < 5.10^-2^ (two-factor ANOVA followed by Tukey’s multiple comparison test of variants versus WT).

### Common polymorphisms associate with Endoglin plasma levels

By meta-analyzing results from two genome wide association studies (GWAS) on Endoglin plasma levels in up to 46,091 healthy individuals, we identified 8 loci significantly (p < 5.10^-8^) associated with Endoglin levels (Figure 4, Supplemental Table 8). The most significant association holds to *ABO* locus (p=4.25.10^-262^), with rs558240 being the lead single nucleotide polymorphism (SNP). The second associated locus mapped to *PLAUR* with the non-synonymous rs4760 being the lead SNP (p=1.14.10^-158^). Remaining loci were *ENG* (rs10987756, p=2.35.10^-43^), *TIRAP* (rs8177398, p=8.82.10^-14^), *NCOA6* (rs73106997, p=1.12.10^-12^), *B3GNT8* (rs284660 p=8.46.10^-10^), *ASGR1* (rs67143157 p=1.83.10^-8^) and *HBS1L* (rs1547247, p=1.98.10^-8^).

**Figure 4.**
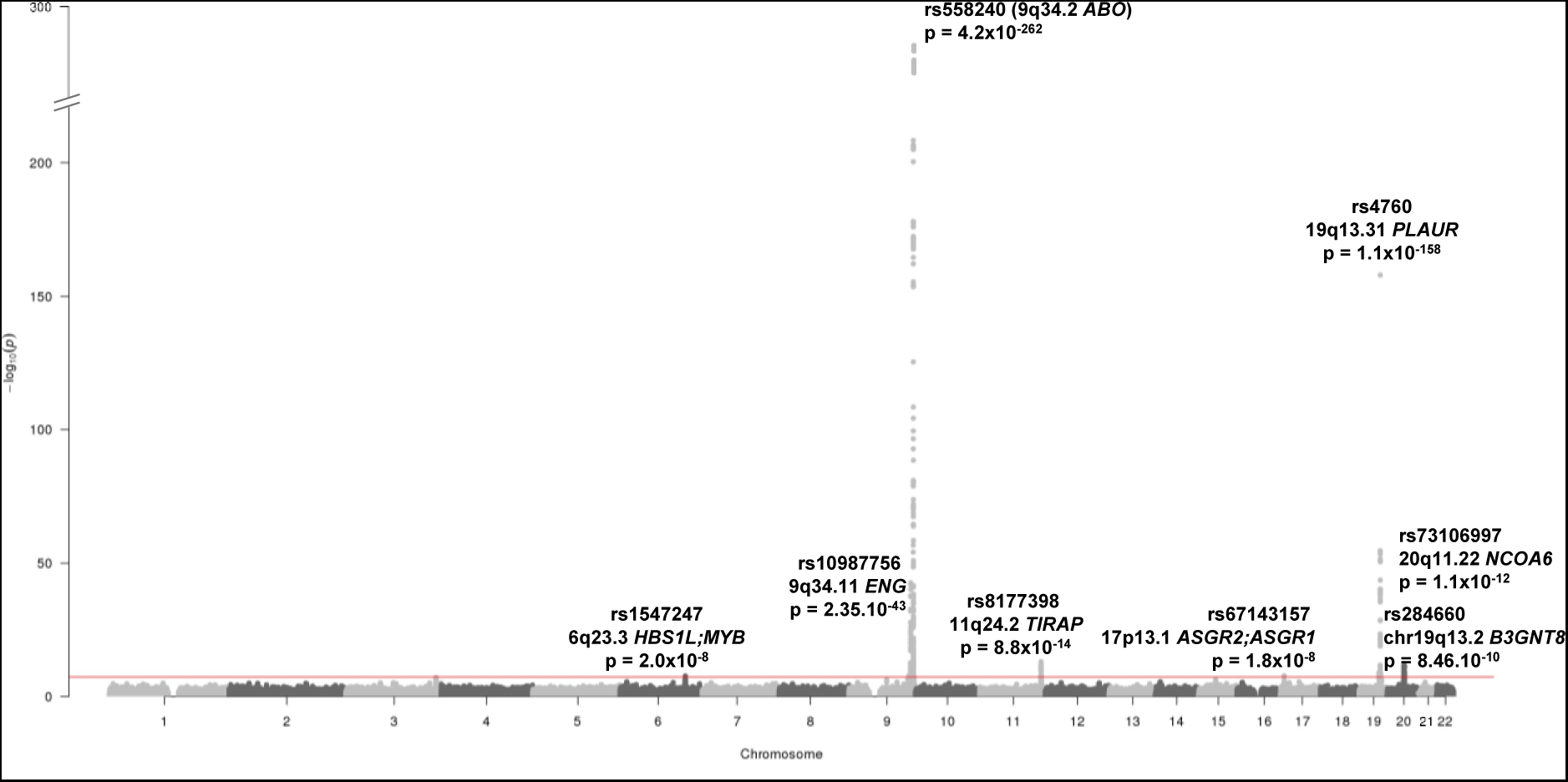
Manhattan plot summarizing the meta-analysis of two GWAS datasets on plasma Endoglin levels in 46,091 individuals. The identity, location and p-value of the 8 loci significantly (p < 5.10^-8^) associated with endoglin levels are indicated.

The *ABO* locus codes for the ABO blood groups whose main groups can be genetically characterized by rs2519093 (A1), rs1053878 (A2), rs8176743 (B), rs8176719 (O1) and rs41302905 (O2)^38^. To further clarify the association observed at the *ABO* locus, we investigated the association of ABO blood groups with Endoglin plasma levels in 966 heathy individuals of the 3C study^39^. The pattern of associations is shown in Figure 5. To summarize, in this healthy population, assuming additive allele effects, the ABO B group was associated with increased Endoglin levels (β = +0.50 ± 0.088, p=1.8.10^-8^) compared to the O1 blood group whereas the A1 group was associated with decreased levels (β = -0.23 ± 0.056, p=5.12.10^-5^). Altogether, ABO blood groups explained 5.8% of the inter-individual variability in Endoglin plasma levels. An additional 1.8% was explained by the 7 lead SNPs at the other genome-wide significant loci.

**Figure 5.**
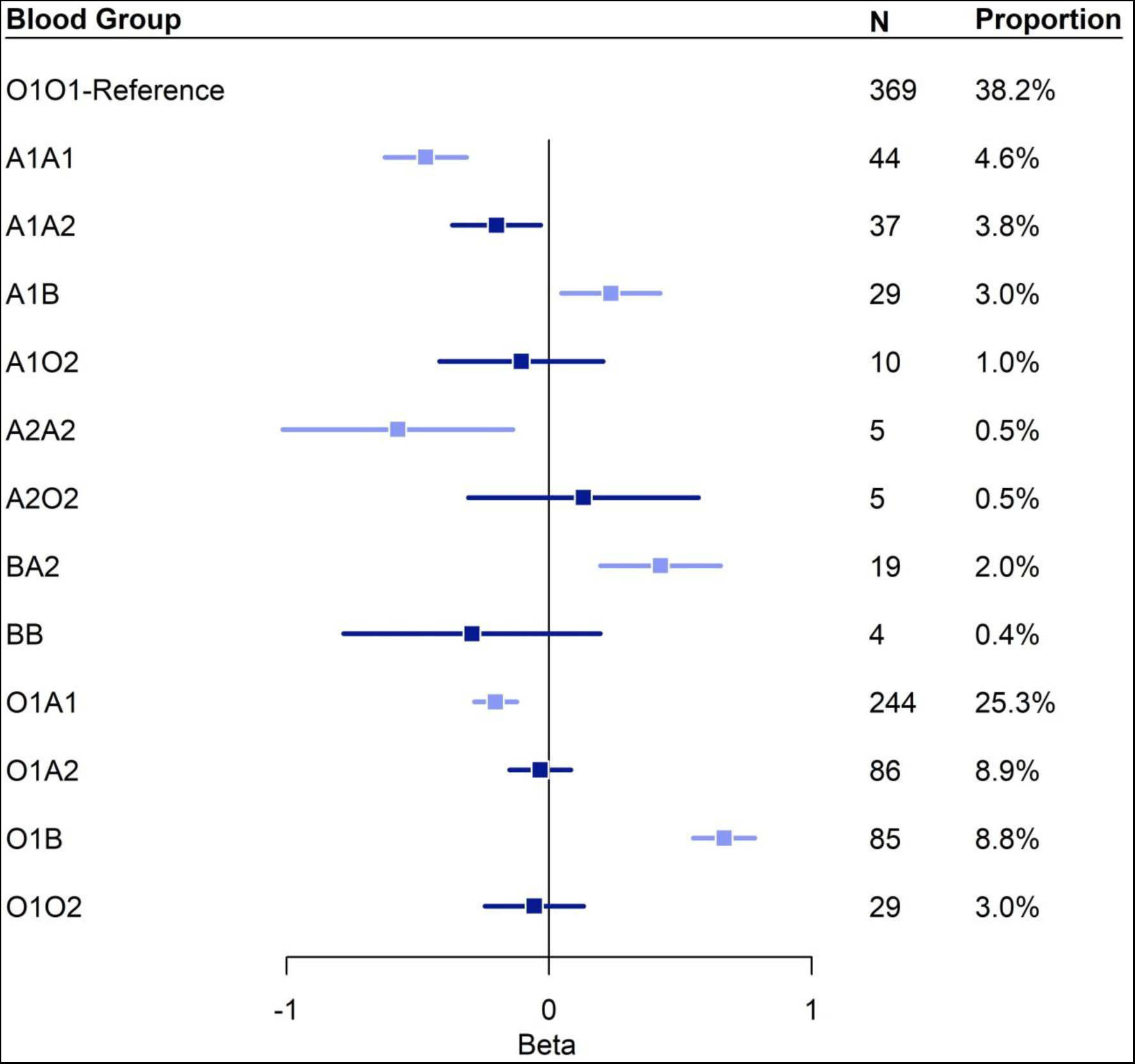
Forest plot summarizing the association of ABO diplotypes with Endoglin plasma levels. Diplotype effects were assessed with the O1O1 diplotype as the reference and were estimated based on standardized Endoglin values measured in 966 participants from the 3C study, where Endoglin levels were determined using Olink technology. N, number of carriers. Association of ABO diplotypes with Endoglin plasma levels were compatible with the additive effects of the B (β = +0.50 ± 0.088, p=1.8.10^-8^) and A1 haplotypes (β = -0.23 ± 0.056, p=5.12.10^-5^).

It is worth noting that the aforementioned *ASGR1* rs67143157 also associated with plasma levels of urokinase-type plasminogen activator receptor (uPAR), encoded by *PLAUR*, both in the deCODE (p=5.64.10^-16^) and in the Fenland (p=4.22.10^-5^) studies. We further assessed the biological correlation between uPAR and Endoglin plasma levels in the 3C study. After adjusting for age and sex, both protein plasma levels strongly correlated with each other (ρ= +0.37, p=2.18.10^-33^). Consistently, the genetic correlation estimated between these two proteins as derived from summary GWAS statistics was ρ= +0.24 (p <10^-16^) (ρ= +0.54 and ρ = +0.15 in Fenland and deCODE, respectively). Of note, in the 3C study, the *ASGR1* rs67143157-G allele also associates with increased uPAR levels (β= 0.08 ± 0.02, p=2.53.10^-4^).

### Association between Endoglin and uPAR in endothelial cells

To follow up on epidemiological findings observed in the 3C study, we investigated the relationship between uPAR and Endoglinin endothelial cells where Endoglin plays a major role. First, we treated human umbilical vein endothelial cells (HUVECs) with recombinant uPAR and observed an increase of Endoglin levels in the supernatant *in vitro* (Figure 6a; Supplemental Table 9). These results strongly suggest that the soluble form of Endoglin could be influenced by uPAR and confirm results from proteogenomics analysis. In order to investigate if intracellular forms of these proteins could also be linked, we performed a knockdown of PLAUR in endothelial cells and observed (p=0.044) for a moderate decrease of Endoglin levels (Figure 6b; Supplemental Table 9), suggesting that also intracellular Endoglin levels can be influenced by uPAR. No significant decrease in ENG RNA levels was obtained in HUVECs with PLAUR knockdown.

**Figure 6.**
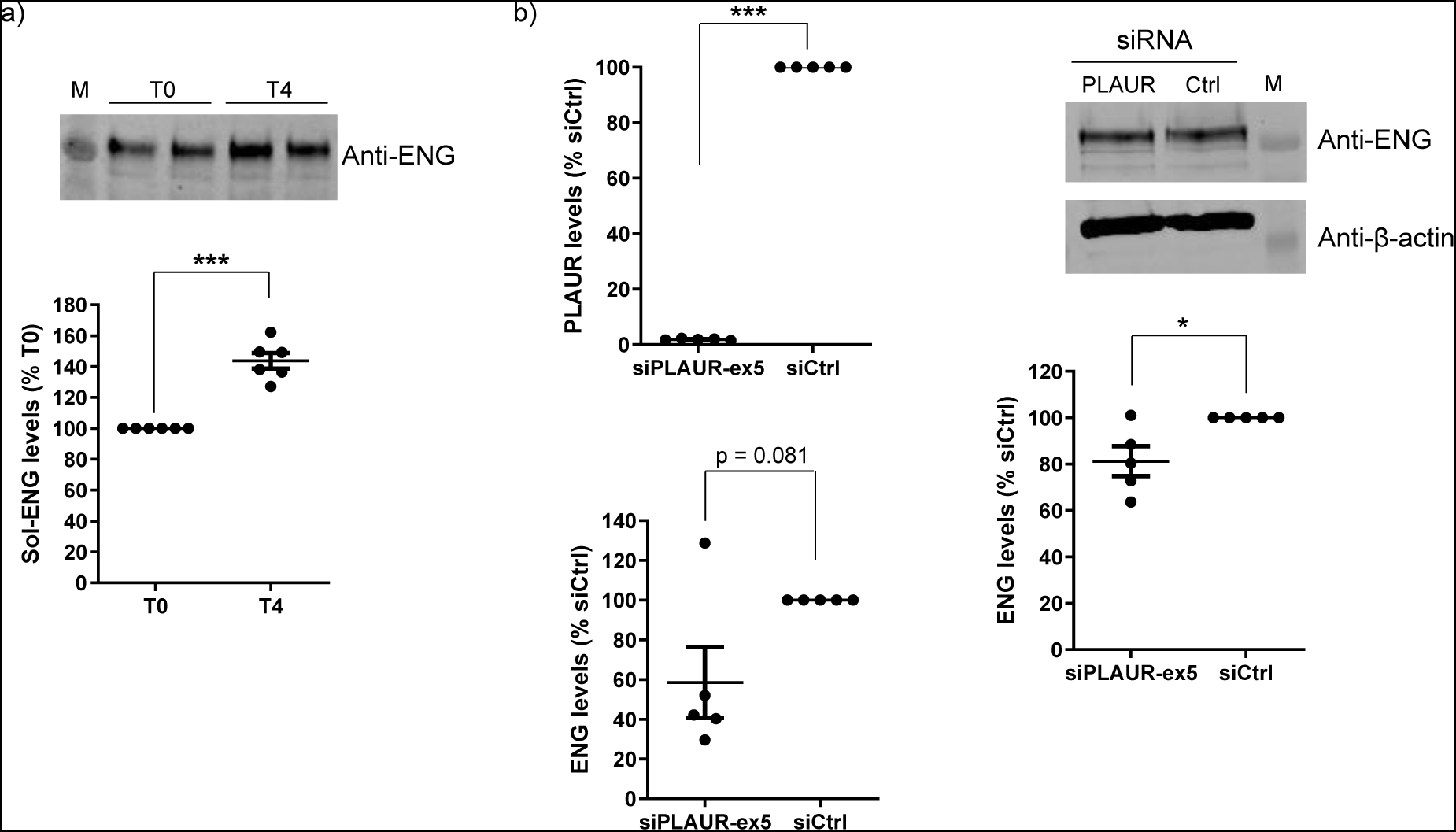
Experimental assays confirm the link between uPAR and Endoglin in endothelial cells in both soluble and intracellular forms. a) Stimulation of HUVEC cells with recombinant uPAR showed an increase in soluble ENG. HUVEC cells were plated in 6 well plates in full medium then in 1 ml of optimem per well overnight. Wells were separately treated with 1 µl of PBS overnight or with 100 ng/ml of recombinant uPAR for 4 hours before recovering the supernatant. Supernatants were treated on 10% western blot gel in order to evaluate soluble ENG levels as shown on the Figure. Quantified data showed an increase of soluble ENG in cells treated with recombinant uPAR compared to those treated with PBS. T0, treatment with PBS overnight; T4, 4 hours uPAR treatment. ENG levels for each sample have been normalized to the total amount of proteins (stain free) then to T0 wells. At least two bands were obtained for the Endoglin, corresponding to the differentially glycosylated Endoglin monomers and they were taken together for the quantification. Graph with standard error of the mean is representative of 6 independent experiments. ***, p-value < 10^-3^, (two-factor ANOVA test of T4 versus T0. b) Knockdown of PLAUR is associated with a modest decrease in intracellular ENG in endothelial cells. HUVECs were plated in 6 well plates and transfected with a PLAUR-specific siRNA in parallel with a control siRNA. Forty-eight hours after transfection, cells were harvested to extract total proteins or RNAs. Normalized 2^-ΔΔCT^ to the siCtrl are shown on the left showing a drastic decrease in PLAUR levels in presence of the siRNA. On the right panel, Endoglin levels for each sample have been normalized to the corresponding β-actin levels then to the siCtrl (%). At least two bands were obtained for the Endoglin, corresponding to the differentially glycosylated Endoglin monomers and they were taken together for the quantification. Graphs with standard error of the mean are representative of 5 independent experiments. ***, p-value < 10^-3^, *, p < 5.10^-2^ (two-factor ANOVA test of siPLAUR-ex5 versus siCtrl).

### Genetic support for a role of soluble Endoglin in human diseases

Building on the GWAS results for plasma Endoglin levels, we conducted Mendelian Randomization (MR) analyses to explore potential causal associations between increased Endoglin levels and various human diseases where soluble Endoglin has been proposed as a biomarker. These diseases correspond to preeclampsia^40^, systemic sclerosis^41^, myocardial Infarction^42^, coronary artery disease^43^ and thrombotic disorders^44^. Statistical evidence for causal association of Endoglin was observed only with venous thrombosis as well as with coronary artery disease after elimination of outliers detected by MR-PRESSO (see methods) (Supplementary Table 10). Finally, these MR results were no longer significant after excluding ABO locus suggesting that these MR findings were partially driven by ABO, a well-known thrombosis-associated locus with strong pleiotropic effects.

## Discussion

The unresolved molecular origins in some HHT cases and the unexplained variability in clinical phenotypes present significant challenges for the diagnosis and management of HHT patients and their families. We here combine the exhaustive characterization of 5’UTR variants altering upORFs in *ENG* and a meta-analysis of GWAS on plasma ENG levels to facilitate the identification of new rare variants in molecular diagnosis of HHT and to bring new molecular elements that could help explaining phenotype variability.

Our group, along with others, recently shed light on a specific class of 5’UTR *ENG* variants that act as LoF variants leading to Endoglin deficiency by identifying in HHT patients 5 variants creating uAUGs at the origin of uoORFs terminating at the same c.125 position in the CDS^14–19^. We here extend these observations by bioinformatically characterizing all 5’UTR *ENG* variants that can create or modify upORFs, aiming to facilitate the identification of pathogenic variants for HHT. This was achieved by cataloguing all possible SNVs (already reported or yet unreported artificial ones) that can create canonical or non-canonical TIS, create new stop codons or delete existing stop codon in the *ENG*. The catalogued variants (Supplemental Table 2) can be quickly queried to identify candidate 5’UTR *ENG* variants in HHT, contributing to resolve molecular origins of HHT. For instance, among *ENG* variants reported in ClinVar, 43 (∼14%) are located in the 5’UTR with 18 (∼42% of 5’UTR variants) annotated with MORFEE as creating uTIS.

By focusing on uAUG creating variants at the origin of uoORFs ending with the uStop-c.125, we experimentally demonstrate that most of these variants are associated with decreased Endoglin levels *in vitro*. We also show that prediction metrics dedicated to upORF-altering variants (Kozak strength, KSS and PreTIS scores ; see Supplemental results) deserve to be evaluated with a larger dataset of variants with different impacts (increase, decrease or null effect) on protein levels. Of note, one of the tested uAUG-creating variants, c.-37G>T, is associated with higher levels of Endoglin in our assay. In addition to the creation of a uAUG, this variant is also predicted to create a stop codon (uUGA). The predicted stop codon is in frame with several non-canonical uTIS (Supplemental Table 10). Noteworthy, 2 of these in frame uTIS (c.-139 and c.-109 CUGs) have been reported in databases to initiate natural uORFs (Supplemental Figure 1a). Consequently, the c.-37G>T could shorten these two natural uORFs. Of note, the one at position c.-139 is encompassed with strong Kozak sequence and has high KSS and PreTIS scores (Supplemental Table 11). Whether this uORF modification could explain the increase of protein levels still need to be explored. This specific case illustrates the complexity to predict the potential impact of variants with multiple consequences on upORF alterations. In addition, we demonstrate, for the first time, the functional impact of a 5’UTR variant that creates a non-canonical uTIS (uCUG) and that associates with reduced levels of ENG *in vitro*. This variant was identified in 2 patients with suspected HHT. These findings suggest that rare *ENG* variants predicted to create non-canonical uTIS in frame with the uStop-c.125 should be considered as candidates for causing HHT.

HHT is characterized by a significant clinical heterogeneity which has been proposed to be attributable, at least partially, to either a second hit variant in HHT genes or to common variants in some modifier genes^6^. In relation to the first scenario, our findings suggest that variations altering upORFs in the 5’UTR of ENG (and potentially in other HHT genes) with strong or moderate effects could be good candidates to explain such heterogeneity. Regarding the second scenario, some works have proposed that the soluble form of Endoglin could also be a relevant biomarker for HHT^25,26^. In that context, our identification of 8 loci presenting common polymorphisms associated with plasma Endoglin levels can also provide new insights about novel molecular players contributing to HHT heterogeneity. We first observe that the *ABO* locus accounts for ∼6% of inter-individual variability. This suggests that the clinical utility of ABO blood groups in the context of clinical diagnosis for HHT, especially for explaining incomplete penetrance of identified (likely) pathogenic *ENG* variations or those associated with moderate effect, deserves further investigations. Our proteomic analyses also pinpoint to a novel association between uPAR and Endoglin in plasma. We further explored this association in endothelial cells and observed that not only soluble uPAR and Endoglin can be associated, but also that intracellular Endoglin levels could be influenced by uPAR. While this association has not been described before, this original finding is supported by reports demonstrating a link betwen Endoglin and urokinase-type plasminogen activator (uPA, uPAR ligand) inhibitor-1 (PAI-1) in cancer associated fibroblasts^23^ and endothelial cells^45^. PAI-1, uPA and uPAR are known to interact together to modulate angiogenesis and fibrinolysis^46,47^. Additional extensive work is needed to deeper characterize the role of PAI-1/uPAR in Endoglin function. Similarly, further studies are mandatory to assess the clinical utility of common genetic variants at the *ABO*, *ASGR1*, *B3GNT8*, *ENG*, *HBS1L*, *NCOA6*, *PLAUR*, and *TIRAP* loci we here identify as modulating plasma Endoglin levels.

Finally, our proteogenomic findings open new therapeutics perspectives. For instance, uPAR has been reported as a pharmaceutical target for Ruxolitinib (a family member of JAK inhibitor) in the context of myelofibrosis and essential thrombocythemia in the Therapeutic Target Database^48^. More recently, uPAR was used as a target of chimeric antigen receptor CAR T cells to improve aging conditions by the elimination of senescent cells^49^. Similarly, *ASGR1* is currently studied as a pharmaceutical target for AMG 529 in the context of diseases of the circulatory system including essential hypertension (https://clinicaltrials.gov/study/NCT03170193).

To conclude, our work provides new insights into the interpretation of *ENG* non-coding variants. It has direct implication in HHT and can also contribute to better understand the implication of ENG in other human conditions. Besides, our plasma proteogenomics investigation coupled with experimental validation identifies the uPAR pathway as a novel regulator of ENG biology warranting further research.

## Methods

### Nomenclature

DNA sequence variant nomenclature follows current recommendations of the HGVS^50^.

### Search for all upORF-altering variants in the 5’UTR of *ENG*

We *in silico* mutated each position in the 5’UTR of the main transcript of ENG (MANE select, ENST00000373203.9) reported in the latest version of Ensembl database (GRCh38.p14) with the 3 alternative nucleotides to generate a vcf file containing all possible SNVs between positions c.-303 (i.e., first nucleotide of the 5’UTR) and c.-1 (Supplemental Table 1). The generated vcf file was then annotated using an updated version of the MORFEE bioinformatics tool^14,51^ now available on https://github.com/CarolineMeg/MORFEE. This version can annotate variations predicted to (i) create canonical and non-canonical TIS; (ii) create new stop codons (TAA, TAG and TGA); and/or (iii) delete existing stop codons along a given transcript. The resulting list of SNVs is provided in Supplemental Table 2.

Additionally, we extracted known upORFs in the 5’UTR of *ENG* from public databases reporting small ORFs that have been identified through ribosome profiling and/or mass spectrometry in human cells: sORFs repository (www.sORFs.org)^35^ ; metamORF database (https://metamorf.hb.univ-amu.fr/)^36^ ; vUTR interface (https://vutr.rarediseasegenomics.org/) ; and smoRFs browser (https://smorfs.ddnetbio.com)^37^.

### Selection of *ENG* variants for experimental validation

Five rare variants creating uAUGs in frame with the stop codon at position c.125 have been previously identified in HHT patients and have shown drastic effects on ENG protein levels^14^. We here selected all additional 8 variations identified by MORFEE to create uAUGs in frame with the stop codon at position c.125 for experimental validation (Supplemental Table 2).

We also selected the c.-76C>T variation, creating a non-canonical uTIS in frame with the c.125 stop codon. This variation was further identified in a collection of HHT patients with unresolved molecular diagnosis (Supplemental Table 2).

### Plasmid constructs

Preparation of pcDNA3.1-L-ENG constructions was performed by directed mutagenesis on pcDNA3.1-L-ENG-WT construct^14^ using the 2-step overlap extension PCR method^52^ and primers listed in Supplemental Table 11. BamHI and SacII or BamHI and BlpI were used as cloning sites, depending on the inserted variant (Supplemental Table 11). Only pcDNA3.1-L-ENG-c.-287C>A construction was prepared by a simple PCR reaction with a forward primer carrying the variant downstream of BamHI cloning site and a reverse primer overlapping SacII (Supplemental Table 12). All new constructs were verified by Sanger sequencing of the insert and cloning sites (Azenta/Genewiz).

### Functional analysis of *ENG* 5’UTR variants

Transfection of HeLa cells, RNA and protein extractions as well as western blot and RT-qPCR analyses were carried out as described in Soukarieh *et al*., 2023^14^. Reverse transcription was performed by using oligo-dT and random hexamers. Primers are listed in Supplemental Table 12.

### Kozak sequence interpretation and bioinformatics predictions

We here used the obtained results in our functional assays on *ENG* variants (Table 1) to evaluate the predictive power of the strength of the Kozak sequence and of 2 predictive scores, TIS-predictor^53^ (KSS scores) and PreTIS^54^ tools.

The Kozak sequence is defined as the genomic sequence surrounding a TIS. The optimal Kozak sequence is [**A/G**]CC<u>ATG</u>**G**, underlined nucleotides corresponding to the TIS and bolded nucleotides to the most conserved positions. We have arbitrarily considered a given Kozak sequence as (i) strong when it contains a purine at position -3 <u>and</u> a guanine at position +4; (ii) moderate when it contains a purine at position -3 <u>or</u> a guanine at position +4, and; (iii) weak when it does not contain a purine at position -3 nor a guanine at position +4.

### Common polymorphisms associated with Endoglin levels

To identify common polymorphisms associated with plasma Endoglin levels, we meta-analyzed GWAS summary statistics from 2 large scale proteogenomics resources where Endoglin has been measured. These resources include 10,708 participants from the Fenland study^55^ and 35,559 Icelander participants of the deCODE project^56^ with Endoglin plasma levels measured using the Somalogic platform. The meta-analysis of these GWAS results was performed using the METAL software implementing the Z-score fixed-effect model^57^. The heterogeneity of genetic associations across studies was assessed using the Cochran-Mantel-Haenszel method and its magnitude was expressed in terms of I^2^ ^58^. Only associations with moderate heterogeneity I^2^ <30% were considered.

To fine map genomic findings obtained from this meta-analysis, we used individual data from an additional plasma proteogenomic resource consisting of a sample of 1,056 population-based participants of the 3C-Dijon study^39^ profiled on the Olink Explore 3072 panel (Supplemental Methods). After exclusion of 3 participants presenting with extremely low outlier Endoglin values, 966 3C-Dijon participants with GWAS data remained for fine mapping analysis. In 3C-Dijon, association of common polymorphisms with Endoglin levels was conducted on centered and standardized NPX values adjusted for age and sex.

### Mendelian Randomization

Capitalizing on the GWAS results on plasma Endoglin levels, we deployed two-sample MR^59,60^ analyses to assess a possible causal role of plasma Endoglin on preeclampsia^40^, systemic sclerosis^41^, coronary artery disease^43^, myocardial infarction^42^ and venous thrombosis^44^ using dedicated summary GWAS statistics. Several MR methodologies were deployed including Inverse Variance Weighted (IVW)^61^, Weighted Median^62^, Egger^63^, and MR-PRESSO^64^ implemented in the TwoSampleMR R package (version 0.5.7) and MR-PRESSO R package (version 1.0). For these MR analyses, we first selected genetic variants that were present in both the Fenland and deCODE GWAS results for plasma ENG levels and showed genome-wide significance (p < 5.10^-8^) in the meta-analysis of both GWAS results and a moderate heterogeneity (I^2^<50%). We then kept as instrument variables genetic variants that remained after clumping for linkage disequilibrium (LD) at r^2^ < 0.01 for a distance of 10Mb (based on European 1000 Genomes phase 3 version 5 reference panel).

### Genetic correlation between plasma Endoglin and uPAR levels

To estimate the correlation between genetically determined plasma levels of Endoglin and of uPAR (urokinase-type plasminogen activator receptor, encoded by *PLAUR*) identified from the aforementioned plasma proteogenomics investigations, we used the Linkage Disequilibrium Score regression approach^65,66^ implemented in the LDSC package (https://github.com/bulik/ldsc). This method was applied to Endoglin and uPAR summary GWAS statistics separately from the Fenland and Decode studies. The Fisher z-transformation^67^ was then applied to the obtained genetic correlation estimates. The two resulting z coefficients were then meta-analyzed using the fixed effect Mantel-Haenszel methodology^58^ and the combined z coefficient was transformed back to obtain a combined estimate of the genetic correlation between plasma levels of Endoglin and uPAR.

### Experimental validation of the association between soluble uPAR and Endoglin

In order to study the potential association between soluble uPAR and sol-ENG, HUVECs were treated with 100 ng/ml of recombinant uPAR (R&D systems). Briefly, HUVECs were seeded in P6 well plates in EGM2 medium from Lonza. At 80% of cell confluence, the medium was replaced with 1 ml of optimem per well and wells were separately treated with 1 µl of PBS overnight or with 100 ng/ml uPAR during 4 hours. Proteins and RNAs were then collected and treated by Western blot and RT-qPCR (Supplemental Table 10) as described above. Supernatants were also collected and equal volumes were treated by western blot with a liquid transfer on PVDF membranes. Sol-ENG was then quantified and normalized to total protein amounts. Total protein labeling was performed using No-Stain protein labeling Reagent (Invitrogen) on membrane (post-transfer). Proteins were visualized using Imager E-BOX Vilber on UV light transilluminator and quantified by densitometry using Fiji-ImageJ software to perform total protein normalization.

### Knockdown of uPAR in HUVEC cells

Intracellular association between uPAR and ENG was also investigated in HUVEC cells. For this purpose, cells were transfected with 30 pmol of a siRNA targeting PLAUR (siPLAUR_ex5: CCAAUGGUUUCCACAACAA)^68^ in parallel with a control siRNA (Eurogentec, SR-CL000-005) by using Lipofectamine RNAiMAX (Invitrogen) following manufacturer’s instructions. Forty-eight hours after transfection, proteins and RNAs were collected and treated by Western blot and RT-qPCR (Supplemental Table 12) as described above.

### Statistical analysis of *in vitro* data

Differential protein and RNA levels were assessed using analysis of variance followed by Tukey’s multiple comparison test when appropriate. A p < 0.05 was used to declare statistical significance.

## Acknowledgments

This project was carried out in the framework of the French National Research Agency (ANR) ANR-23-CE17-0042-01 program as part of the ENDOMORF project and of the INSERM GOLD Cross-Cutting program (D-A.T.). O.S was financially supported by a grant of the Lefoulon-Delalande Foundation. IC was supported by the Digital Public Health Graduate Program (DPH), a PhD program supported by the French Investment for the Future Program (grant no. 17-EURE-0019). The 3C proteomics project was supported by a grant overseen by the French National Research Agency (ANR) as part of the “Investment for the Future Programme” ANR-18-RHUS-0002 and by the Precision and global vascular brain health institute funded by the France 2030 investment plan as part of the IHU3 initiative under grant agreement ANR-23-IAHU-0001. Statistical analyses benefited from the CBiB computing centre of the University of Bordeaux.

## Author contributions

OS and DAT conceived the project. OS, CP and BJV designed the experiments. OS, CP, CD, and BJV performed the experiments. OS, CP, CD, and BJV analyzed the data. BJV and AG provided technical support and suggestions on the project and the experiments. SDG and MT were in charge of clinical management of HHT patients. CM performed the mutational saturation and variant annotation with MORFEE. GM analyzed proteogenomics data. IC performed bioinformatics analysis on the 3C study under the supervision of SD. OS and DAT drafted the paper that was further shared to co-authors who read/corrected/ and approved the final manuscript.

## Competing interests

The authors declare that they have no known competing financial or non-financial interests or personal relationships that could have appeared to influence the work reported in this paper.

## Data availability

ENG constructs generated during this study are available upon request by email from the corresponding author.

## Code Availability

The used version of MORFEE tool is available at https://github.com/CarolineMeg/MORFEE.

## Notes

### Competing Interest Statement

The authors have declared no competing interest.

### Author Declarations

These resources include 10,708 participants from the Fenland study, 35,559 Icelander participants of the Decode project, and 1,056 population-based participants of the 3C-Dijon study.

### Summary of Updates

New Figure 6, and its corresponding results and methods. New order of authors. New MR analysis on different diseases. The whole manuscript has been updated but taking into account the new data generated.

