## Supplemental Tables and Figures for "Unveiling Endoglin non canonical regulation: spotlight on the new role of the uPAR pathway"

**Short title: Non-canonical genetic regulation of Endoglin**

Gaëlle Munsch[1], Carole Proust[1], Clémence Deiber[1], Caroline Meguerditchian[1], Ilana Caro[1], Maud Tusseau[2], Alexandre Guilhem[3,4,5], Shirine Mohamed[6], INVENT consortium, Aurélie Goyenvalle[7], Stéphanie Debette[1,8], Béatrice Jaspard-Vinassa[9], Sophie Dupuis-Girod[2,10], David-Alexandre Trégouët[1*], Omar Soukarieh[1,9*]

[1] Univ. Bordeaux, INSERM, Bordeaux Population Health Research Center, UMR 1219, F-33000 Bordeaux, France

[2] Hospices Civils de Lyon, French National HHT Reference Center and Genetics department, Hôpital Femme-Mère-Enfant, 69677, Bron, France

[3] Hospices Civils de Lyon, Service de Génétique, Groupement Hospitalier Est, 69677, Bron, France

[4] Centre de Référence National pour la maladie de Rendu-Osler, Groupement Hospitalier Est, Bron, France

[5] TAI-IT Autoimmunité Unit RIGHT-UMR1098, Université de Bourgogne, INSERM, EFS-BFC, Besançon, France.

[6] Département de Médecine interne et Immunologie Clinique, CHRU BRABOIS, Vandoeuvre-lès-Nancy, France

[7] Université Paris-Saclay, UVSQ, Inserm, END-ICAP, Versailles, France

[8] Department of Neurology, Institute for Neurodegenerative Diseases, Bordeaux University Hospital, France

[9] Univ. Bordeaux, INSERM, Biology of Cardiovascular Diseases, U1034, F-33600 Pessac, France

[10] Univ. Grenoble Alpes, Inserm, CEA, Laboratory Biology of Cancer and Infection, F-38000 Grenoble, France.

### Supplemental Methods

#### Proteomic data analysis in the 3C-Dijon study

Olink proteomic profiling was conducted on EDTA plasma tube of more than 500 µL not thawed/refrozen using the PEA technology following the manufacturer's protocol[1]. Profiling was performed at the McGill Genome Center (Montreal, Canada). Pre-processing of the proteomic data included plate-based normalization and QC checks based on appropriate Olink protocols. Data were transformed and normalized to Olink's Normalized Protein eXpression (NPX) values, relative protein quantification unit in a logarithmic base 2 scale. Twelve samples were removed after principal component analysis of all proteins because they are found to deviate of more than 5 standard deviations from the mean. Besides, three proteins were removed as more than 50% of NPX values were below the protein's detection limit.

### Supplemental Results

#### Bioinformatics predictions partially correlate with the observed functional effects

Among the 13 5'UTR variants we experimentally studied, all leading to uAUGs at the origin of uoORFs ending at stop codon c.125, 6 are characterized by a moderate Kozak sequence (Table 1). These 6 variants (c.-271G>T, c.-249C>G, c.-142A>T, c.-127C>T, c.-33A>G and c.-31G>T) were all associated with decreased protein levels in our assay with 5/6 variants showing the most drastic effects. The 7 remaining variants were associated with a weak Kozak sequence and 5 of them (c.-182C>A, c.-167C>A, c.-79C>T, c.-68G>A and c.-10C>T) were associated with a decrease of the protein levels. Finally, the 2 variants with no decrease of Endoglin protein levels (c.-287C>A and c.-37G>T) are associated with weak Kozak sequences. Our analysis suggests that the sole information on the Kozak sequence of uTIS cannot be used to predict the potential effect of uAUG-creating variants in ENG.

We extended the predictions of the Kozak sequence by applying predictions from TIS-predictor, which takes into account the 10 nucleotides surrounding a given canonical or non-canonical TIS to generate KSS scores reflecting the strength of Kozak sequences. The authors defined a threshold of 0.64 for translation initiation by uAUGs. We extracted KSS scores for our 13 uAUG-creating variants of interest (Table 1; Supplementary Table 2) and found that 6/13 uAUGs have scores higher than 0.64. These 6 variants (c.-271G>T, c.-249C>G, c.-142A>T, c.-127C>T, c.-79C>T and c.-33A>G) were observed to decrease the protein levels of Endoglin in our experiments. Interestingly, these variants are those showing the most drastic effects in our assays. However, 5/11 variants decreasing the protein levels (c.-182C>A, c.-167C>A, c.-68G>A, c.-31G>T and c.-10C>T) presented with KSS scores below the recommended threshold of 0.64 as the two variants with no decrease of protein levels.

Furthermore, we evaluated the efficiency of PreTIS scores to predict uTIS that can alter the protein levels. These scores predict the efficiency of a given uTIS to initiate the translation. We hypothesized that the highest PreTIS scores predicting the most efficient uTIS would be associated with the most drastic effect on the protein levels. We collected PreTIS scores for the 10/13 uAUGs created by 5'UTR variants in ENG. The remaining 3 variants being located within the first 99 nucleotides of the 5'UTR (c.-287C>A, c.-271G>T and c.-249C>G), they cannot be predicted with PreTIS. The totality of the predicted uAUGs (10/10) has PreTIS scores higher than the predefined threshold for translation efficiency (0.54) (Table 1). Importantly, 9 are associated with a decrease of the protein levels in our assays but the remaining one (c.-37G>T) increases Endoglin levels. These observations suggest that while PreTIS could be efficient to predict which uTIS creating variant could have a functional impact, it cannot predict the direction of its molecular effect.

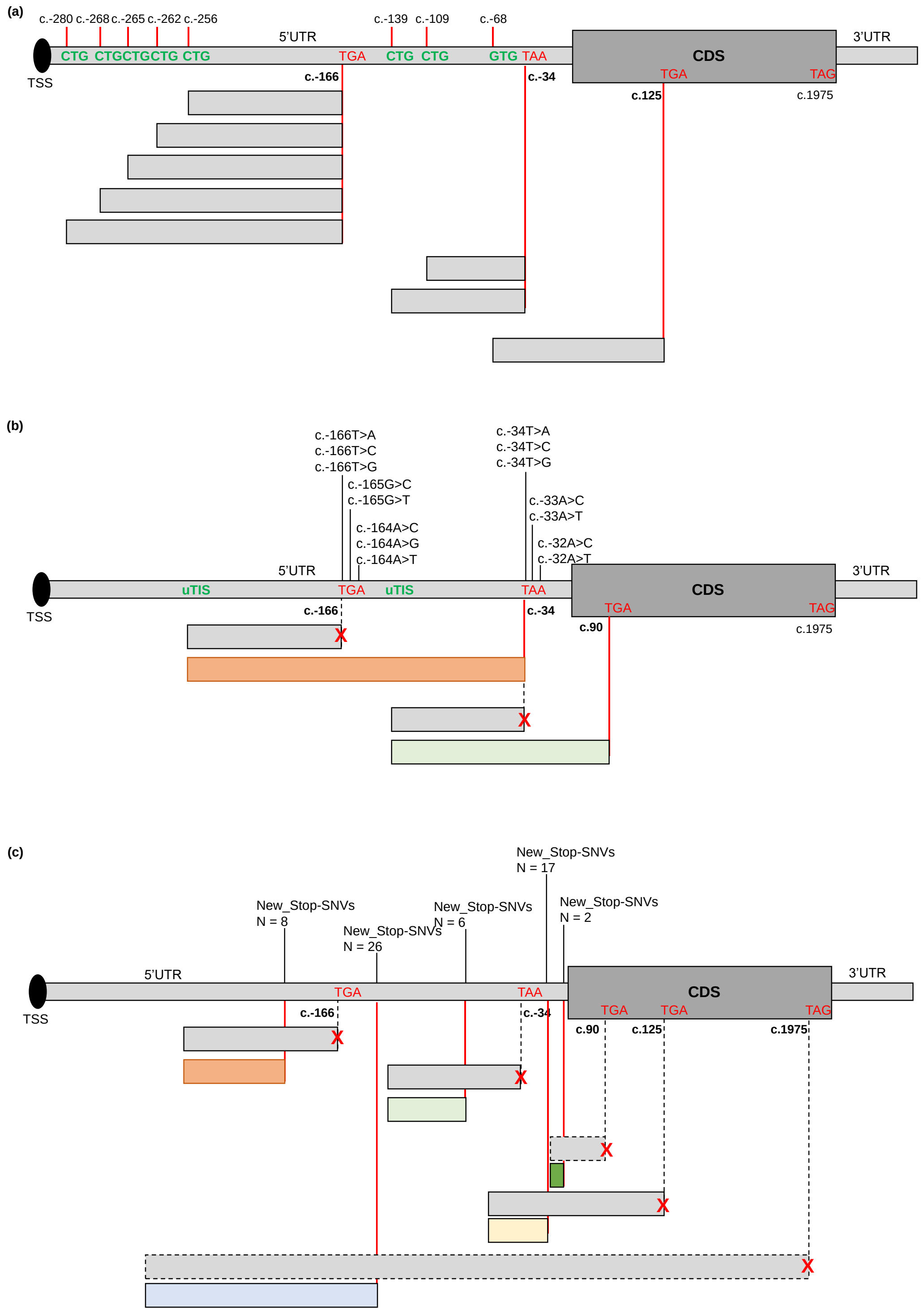


**Supplementary Figure 1. Illustration of naturally existing upstream Open Reading Frames (upORFs) (a) uStop-deleting variants (b) and new-Stop-creating variants in the 5'UTR of ENG.** The type of upstream Translation Initiation Sites (uTIS) and position of stop codons associated with the existing or the new upORFs are indicated. The red cross represents the deleted uStop replaced by another one downstream (b) or the shortening of existing upORFs by the creation of upstream stop codons. The number of variants creating stop codons in the same frame is indicated. TSS, translation start site; CDS, CoDing Sequence; SNVs, single nucleotide variants.

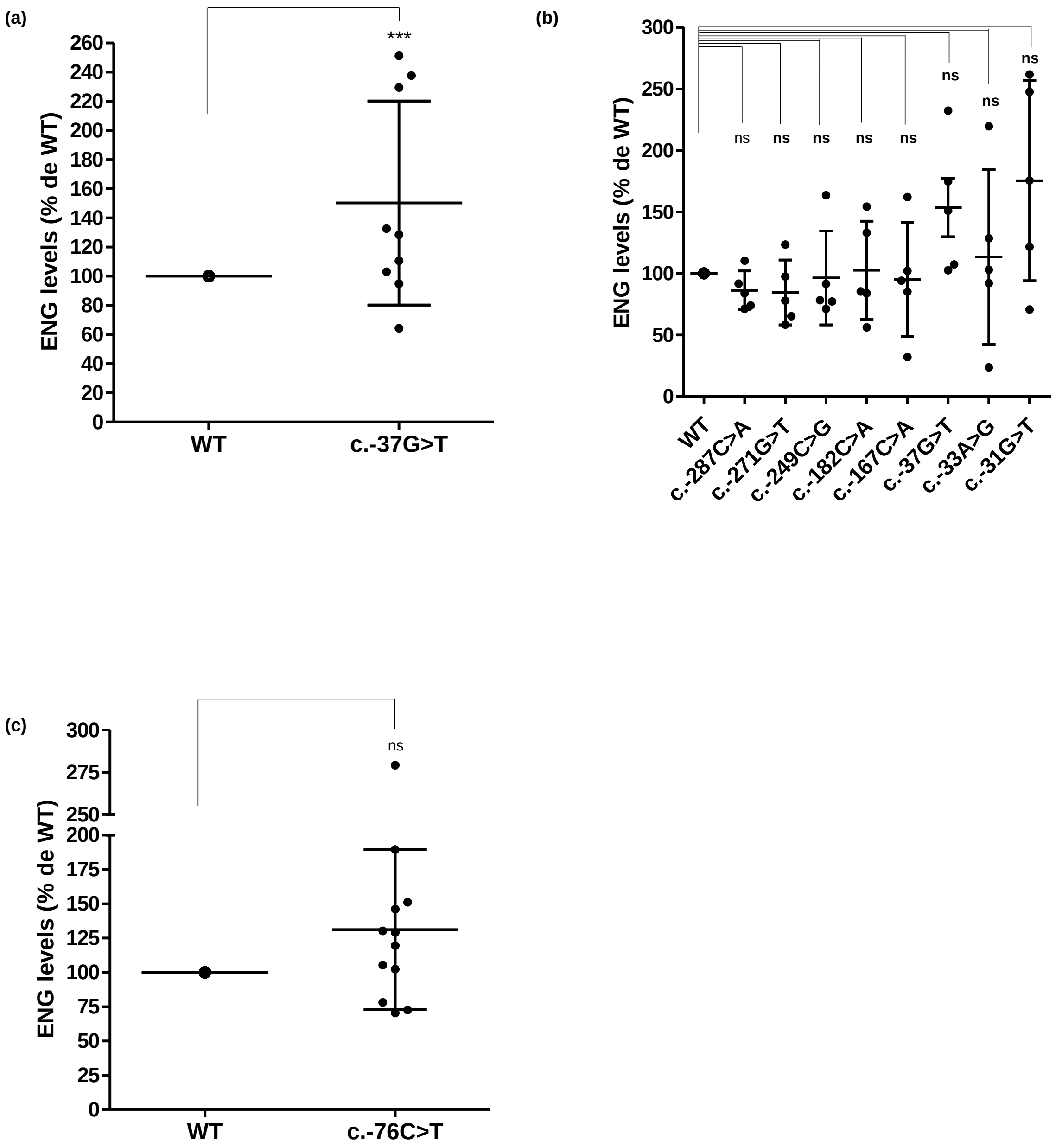


**Supplementary Figure 2. Quantification of ENG levels in HeLa cells from Western blot (a) AND rt-qPCR (c) analyses.** ENG steady-state level in HeLa cells is significantly increased with the c.-37G>T variant in comparison to the wild-type (WT) construct. For quantification, the average of the duplicate has been calculated from the quantified values and ENG levels have been normalized to the corresponding β-actin levels then to the WT (%). The two bands obtained for the Endoglin, corresponding to the more glycosylated (upper band) and less/non glycosylated (lower band) ENG monomers, were taken together for the quantification. The graph with standard error of the mean is representative of 9 independent experiments. (b and c) RNA levels of Endoglin do not variate between wild-type and variants. Normalized 2-ΔΔCT to the WT are shown. Graphs with standard error of the mean are representative of 5 (b) and 12 (c) independent experiments. ***, p-value < $10^{-3}$, ns, non-significant (two-factor ANOVA followed by Tukey's multiple comparison test of variants versus WT).
